# Autoantibodies stabilize neutrophil extracellular traps in COVID-19

**DOI:** 10.1101/2021.03.31.21254692

**Authors:** Yu Zuo, Srilakshmi Yalavarthi, Sherwin Navaz, Claire Hoy, Alyssa Harbaugh, Kelsey Gockman, Melanie Zuo, Jacqueline A. Madison, Hui Shi, Yogendra Kanthi, Jason S. Knight

## Abstract

The release of neutrophil extracellular traps (**NETs**) by hyperactive neutrophils is recognized to play an important role in the thromboinflammatory milieu inherent to severe presentations of COVID-19. At the same time, a variety of functional autoantibodies have been observed in individuals with severe COVID-19 where they likely contribute to immunopathology. Here, we aimed to determine the extent to which autoantibodies might target NETs in COVID-19 and, if detected, to elucidate their potential functions and clinical associations. We measured anti-NET antibodies in 328 individuals hospitalized with COVID-19 alongside 48 healthy controls. We found high anti-NET activity in the IgG and IgM fractions of 27% and 60% of patients, respectively. There was a strong correlation between anti-NET IgG and anti-NET IgM (r=0.4, p<0.0001). Both anti-NET IgG and IgM tracked with high levels of circulating NETs, impaired oxygenation efficiency, and high circulating D-dimer. Furthermore, patients who required mechanical ventilation had a greater burden of anti-NET antibodies than did those not requiring oxygen supplementation. Levels of anti-NET IgG (and to a lesser extent anti-NET IgM) demonstrated an inverse correlation with the efficiency of NET degradation by COVID sera. Furthermore, purified IgG from COVID sera with high levels of anti-NET antibodies impaired the ability of healthy control serum to degrade NETs. In summary, many individuals hospitalized with COVID-19 have anti-NET antibodies, which likely impair NET clearance and may potentiate SARS-CoV-2-mediated thromboinflammation.

## INTRODUCTION

While it has been more than a year since the initial outbreak, coronavirus disease 2019 (**COVID-19**)—caused by the novel severe acute respiratory syndrome coronavirus 2 (**SARS-CoV-2**)—remains a global health challenge with alarming death tolls (1). Many survivors of COVID-19 continue to suffer from post-acute sequelae of the infection and the cause of these long-term symptoms remains for the most part unknown (2-4). Severe, acute COVID-19 is characterized by a thromboinflammatory state driven by a complex interplay between innate and adaptive immune responses (1). This state manifests clinically as acute respiratory distress syndrome, and, in some patients, widespread thrombotic microangiopathy.

Activated neutrophils—and, in particular, neutrophil extracellular traps **(NETs)**—continue to receive significant attention as drivers of SARS-CoV-2-mediated thromboinflammation. NETs are extracellular webs of DNA, histones, and microbicidal proteins released from activated neutrophils via a cell death program called NETosis. Neutrophils presumably deploy NETs to trap and kill pathogens (5); however, NETs may also be key players in the pathophysiology of thromboinflammatory diseases such as cancer, lupus, antiphospholipid syndrome (**APS**), and— based on recent work—COVID-19 (6-8). Indeed, our group and others have described high levels of NETs circulating in the blood of hospitalized COVID patients, where they correlate with disease severity (6, 9-12). We have also found that neutrophil hyperactivity at the time of hospital admission predicts a more severe hospital course (13), and that NET levels are especially high in patients who experience thrombotic complications (14).

Another hallmark of COVID-19 is the development of autoantibodies against a variety of self-antigens, particularly among COVID patients with severe disease (15-18). Many of those autoantibodies appear to perturb normal immune function while influencing disease severity and progression. For example, anti-type I interferon antibodies attenuate a presumably protective immune response against SARS-CoV-2 and thereby exacerbate disease (19). Autoantibodies against annexin A2 and other immunomodulatory proteins are also associated with severe COVID-19 (20, 21). Furthermore, work by our group found that many hospitalized COVID patients developed antiphospholipid antibodies routinely found in APS, an acquired autoimmune thrombophilia (15). Mechanistically, these antibodies promote pathogenic NET formation and accelerate thrombosis *in vivo*.

We recently found high levels of autoantibodies targeting NETs themselves in patients with APS (22), where they impair NET clearance and activate the complement cascade with the potential to amplify thromboinflammation (22). Here, we sought to evaluate the presence of anti-NET antibodies in patients hospitalized with COVID-19 and to determine their potential functions and clinical associations.

## RESULTS

### Measurement of anti-NET activity in COVID-19

Utilizing a unique ELISA platform that we developed (**Figure 1A**), we measured anti-NET IgG and IgM antibodies in 328 patients hospitalized with COVID-19 alongside 48 healthy controls. The clinical characteristics of these patients are described in **Table 1**. Elevated levels of anti-NET IgG and IgM were detected in patients with COVID-19 as compared with healthy controls (**Figure 1B-C**). Based on a threshold set at two standard deviations above the control mean, 89 COVID patients (27%) had high anti-NET IgG activity, while 197 (60%) had high anti-NET IgM activity. We also noted a strong correlation between anti-NET IgG and anti-NET IgM (r=0.4, p<0.0001, **Supplementary Figure 1**). Beyond the ELISA platform, we also assessed anti-NET activity by immunofluorescence microscopy. When paraformaldehyde-fixed NETs were incubated with sera from COVID patients with high levels of anti-NET IgG, antibodies robustly decorated NET strands (**Figure 1D**). We had sufficient sera available to test antiphospholipid antibodies in 171 COVID patients. Positive correlations were noted between anti-NET IgG/IgM and anticardiolipin IgG/IgM and anti-phosphatidylserine/prothrombin IgG/IgM. Anti-NET IgM was also weakly correlated with anti-beta-2 glycoprotein I IgM (**Supplementary table 1**). In summary, elevated levels of anti-NET IgG and IgM antibodies are present in patients hospitalized with COVID-19.

**Figure 1:**
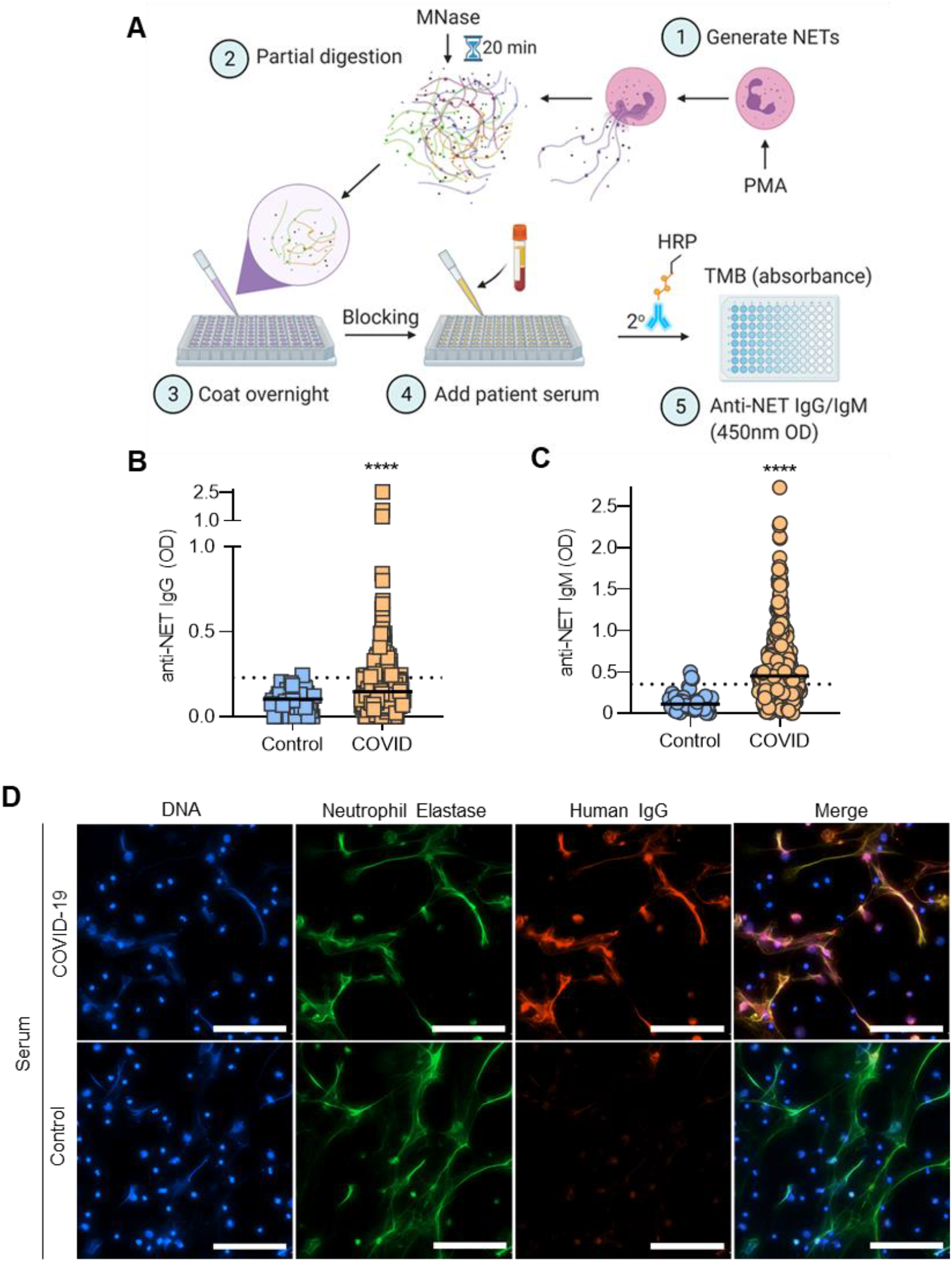
Detection of anti-NET IgG/IgM in sera of COVID patients. **A**, Schematic illustration of anti-NET ELISA. **B-C**, Anti-NET IgG and IgM were measured in sera from 328 hospitalized COVID patients and 48 healthy controls. Levels of anti-NET IgG and IgM at 450-nm optical density (OD) were compared by Mann-Whitney test; ****p<0.0001. Solid lines indicate medians and dotted lines indicate thresholds set at 2 standard deviations above the control mean. **D**, Control neutrophils were stimulated with PMA to generate NETs. Fixed NETs were then incubated with COVID serum with high anti-NET antibodies or healthy control serum; scale bars=100 microns.

**Table 1:** Demographic and clinical characteristics of COVID patients

| **Demographics** |  |  |
| --- | --- | --- |
| Number | 328 |  |
| Age (years)* | 59 ± 17 | (16-95) |
| Female | 139 | (42%) |
| White/Caucasian | 142 | (43%) |
| Black/African-American | 138 | (42%) |
| **Comorbidities** |  |  |
| Diabetes | 130 | (40%) |
| Heart disease | 139 | (42%) |
| Renal disease | 117 | (36%) |
| Lung disease | 134 | (41%) |
| Autoimmune | 13 | (4%) |
| Cancer | 43 | (13%) |
| History of stroke | 22 | (7%) |
| Obesity | 172 | (52%) |
| Hypertension | 193 | (59%) |
| Immune deficiency | 59 | (18%) |
| History of smoking | 82 | (25%) |
| **In-hospital thrombosis** |  |  |
| Arterial thrombosis | 2 | (0.6%) |
| Venous thrombosis | 19 | (6%) |
| Both | 1 | (0.6%) |
| **Final outcome** |  |  |
| Discharged | 261 | (80%) |
| Death | 67 | (20%) |
| * Mean ± standard deviation (range) |  |  |

### Anti-NET antibodies correlate with circulating markers of NET release

Circulating markers of NET release—myeloperoxidase-DNA complexes and calprotectin—were assessed in 171 COVID patients who had sufficient sera available for this analysis. Anti-NET IgG and IgM both demonstrated positive correlations with these markers (**Figure 2A-B**).

**Figure 2:**
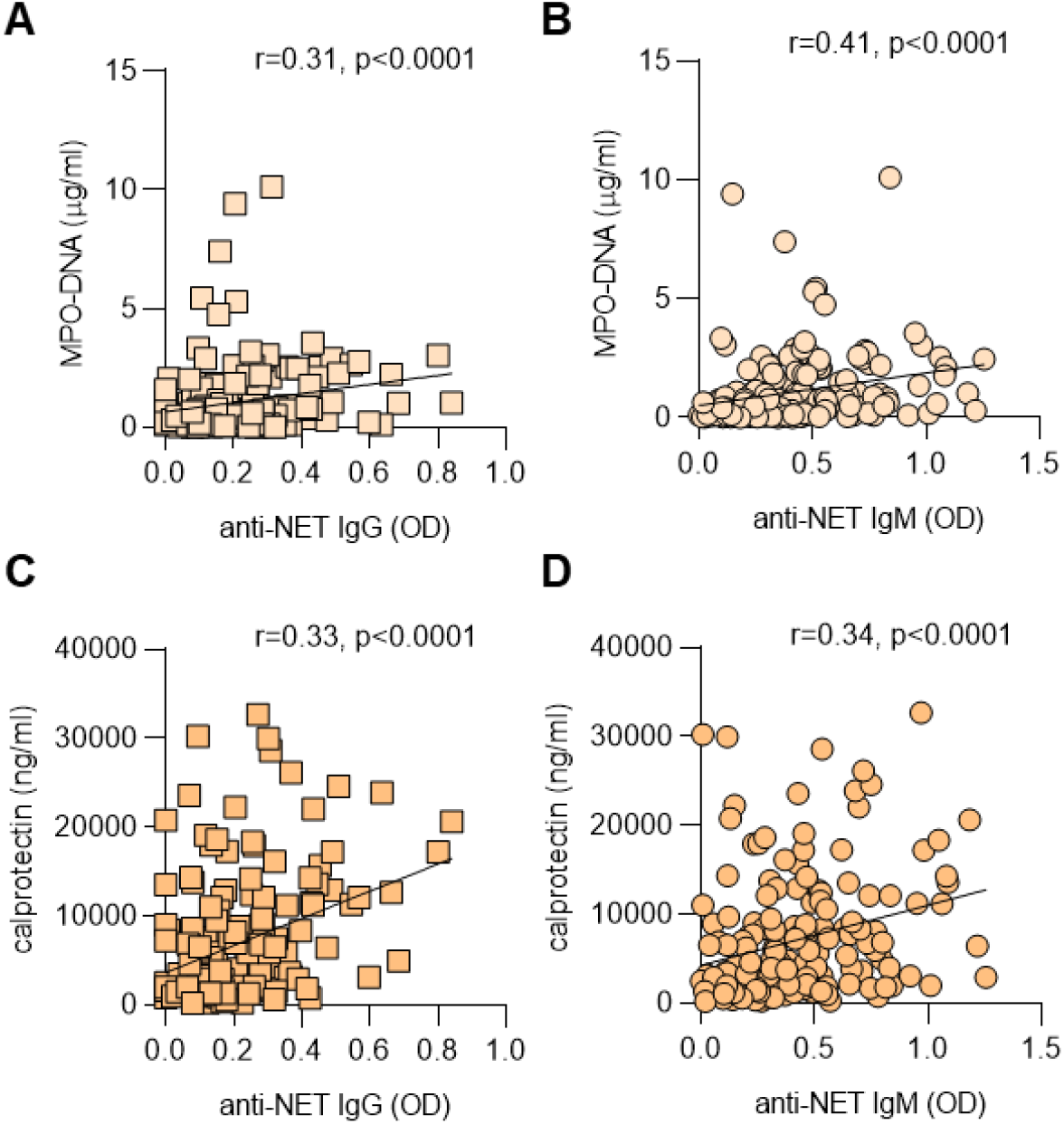
Correlation between anti-NET IgG/IgM and circulating markers of NETs. Spearman’s correlation coefficients were calculated and are shown in the panels (n=171 COVID patients for all panels).

### Association of anti-NET activity with clinical parameters

We next asked whether the presence of anti-NET IgG and IgM associated with various clinical parameters. Specifically, we assessed potential correlations with factors that track with COVID severity including D-dimer (**Figure 3A-B**), SpO_2_ /FiO_2_ ratio (oxygenation efficiency, **Figure 3C-D**), neutrophil count (**Supplementary Figure 2A-B**), and platelet count (**Supplementary Figure 2C-D**). Specifically, anti-NET IgG and IgM both demonstrated positive correlations with D-dimer, neutrophil count, and platelet count, while showing negative correlations with oxygenation efficiency. To determine associated clinical status, we compared serum samples of patients requiring mechanical ventilation (n=140) to patients with oxygen saturation ≥94% on room air (n=69). As compared with patients breathing room air, patients requiring mechanical ventilation had significantly higher levels of anti-NET IgG and IgM (**Figure 3E-F**). In summary, anti-NET IgG and IgM levels track with various measures of COVID severity. Most notably, they are associated with impaired oxygen efficiency and requirement for mechanical ventilation.

**Figure 3:**
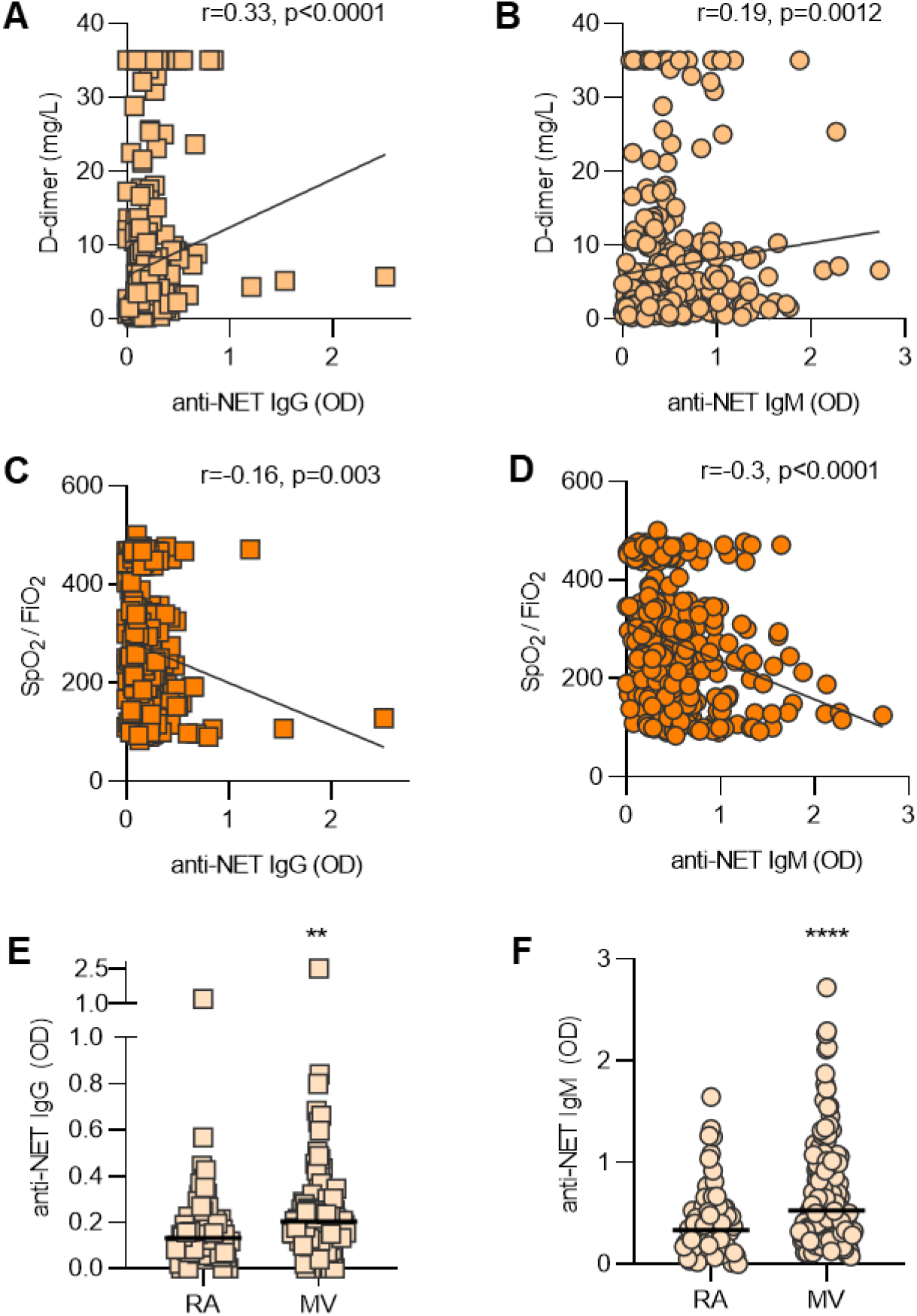
Clinical associations of anti-NET IgG/IgM. **A-D**, Anti-NET IgG and IgM levels were compared to D-dimer (n=287) (A-B) and SpO_2_ /FiO_2_ (n=322) (C-D) on the same day as the COVID-19 research sample. Spearman’s correlation coefficients were calculated and are shown in the panels. **E-F**, COVID patients were grouped based on clinical status: room air (n=69) and mechanical ventilation (n=140). Levels of anti-NET IgG and IgM were compared by Mann-Whitney test; **p<0.01, **p<0.0001. Horizontal lines indicate medians. RA=room air; MV=mechanical ventilation.

### Relationship between anti-NET antibodies and NET degradation

Work by our group and others has revealed that one function of anti-NET antibodies in patients with lupus (23) and APS (22) is to impair NET degradation. Here, we used a NET degradation assay to ask whether COVID sera with high anti-NET activity might demonstrate an impaired ability to degrade NETs (**Figure 4A**). In a cohort of 69 COVID patients with sufficient sera for this analysis, both anti-NET IgG (**Figure 4B**) and anti-NET IgM (**Supplementary Figure 3**) positively correlated with residual NETs after a 90-minute incubation, indicating an impaired ability to degrade NETs. The correlation was relatively stronger for anti-NET IgG (r=0.39, p=0.0009) than for anti-NET IgM (r=0.27, p=0.023). To further confirm the relationship between anti-NET IgG and NET degradation, total IgG was purified from four COVID patients with high anti-NET IgG and supplemented into healthy control serum. These were tested alongside IgG pooled from healthy controls. Control serum supplemented with the COVID IgG demonstrated an impaired ability to degrade NETs as compared with control serum supplemented with control IgG (**Figure 4C**). In summary, high anti-NET activity associated with an impairment in NET degradation by COVID sera.

**Figure 4:**
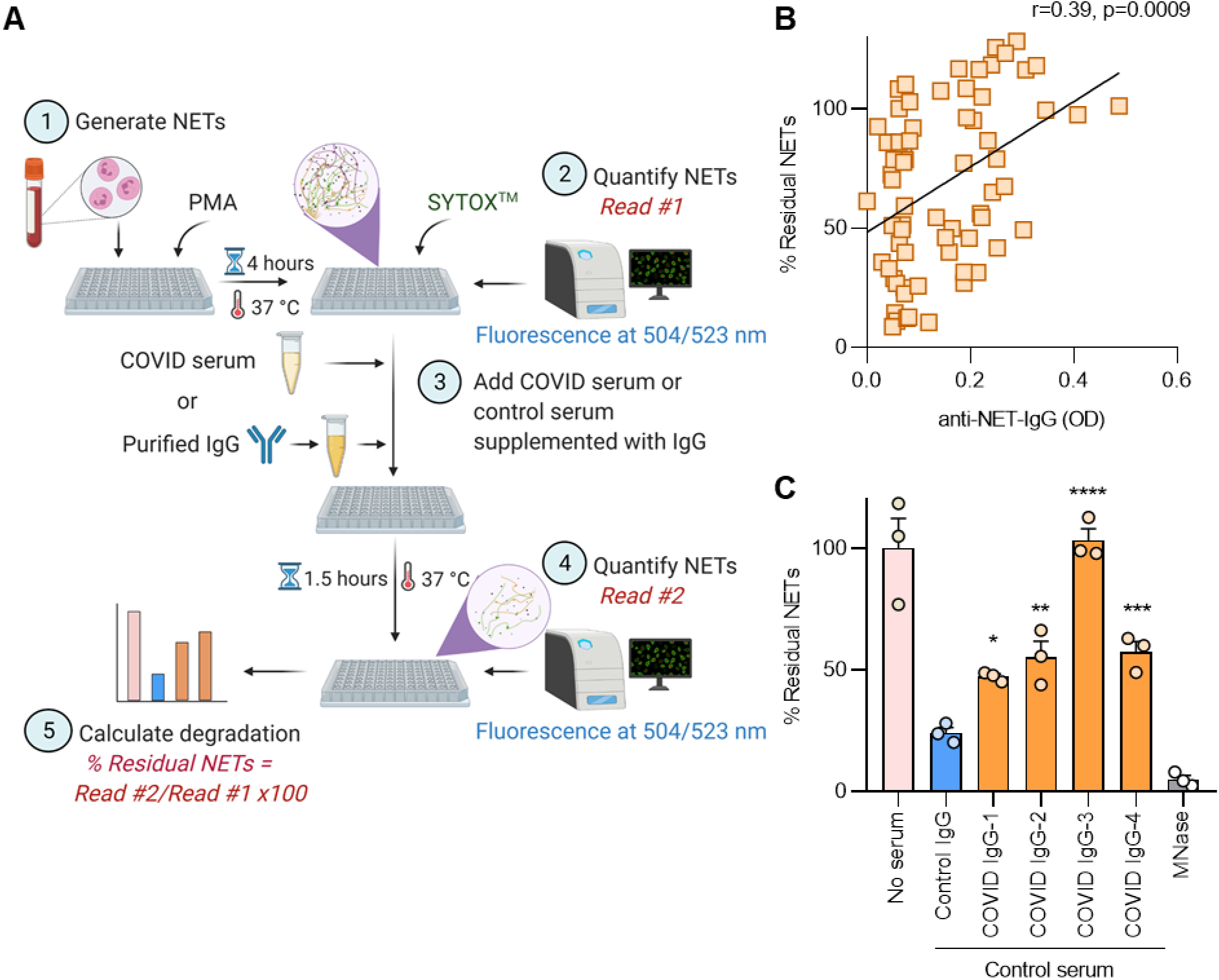
COVID-19 serum and IgG impair NET degradation. **A**, Schematic illustration of NET degradation assay. **B**, Freshly-induced NETs were incubated with COVID-19 serum (n=69). Percent residual NETs was then determined for each sample after 90 minutes. Correlation with anti-NET IgG was determined by Spearman’s method. **C**, Freshly-induced NETs were also incubated with control serum supplemented with either purified IgG from COVID patients or controls, and percent residual NETs was determined for each sample after 90 minutes. Some samples were treated with Micrococcal nuclease as a positive control. Untreated NETs (no serum) acted as negative control. Percent residual NETs was normalized to the mean of untreated NETs. COVID IgG was compared to control by one-way ANOVA with correction for multiple comparisons by Dunnett’s method; *p<0.05, **p<0.01, ***p<0.001, and ****p<0.0001.

## DISCUSSION

In COVID-19, NETs may be directly induced by SARS-CoV-2 (1, 24). They may also be triggered indirectly via activated platelets and prothrombotic autoantibodies (1, 15). Once formed, NETs likely exert direct cytotoxic effects against pulmonary epithelium resulting in alveolar damage and fibrosis (1). They can also injure endothelial cells leading to microvascular damage and thrombotic microangiopathy in lungs, kidneys, and heart (1). Here, we explored the hypothesis that dysfunctional NET clearance may also contribute to COVID pathogenesis.

SARS-CoV-2 appears to have a unique relationship with the immune system. It evades host immune surveillance during early infection leading to high viral loads in some patients (1). As a result, the body then mounts a compensatory hyper-immune response in pursuit of viral clearance. This is characterized by the presence of a lupus-like peripheral B cell compartment in which naïve B cells take an extrafollicular route to becoming antibody-producing cells, and in doing so bypass the normal tolerance checkpoints against autoimmunity provided by the germinal center (25). While this strategy may quickly produce a large amount of virus-neutralizing antibodies, it also sets the stage for the *de novo* production of various pathogenic autoantibodies.

NETs appear to elicit autoantibody production in systemic autoimmune diseases such as lupus, rheumatoid arthritis, and ANCA-associated vasculitis (26). For example, it has been suggested that increased NET formation, the presence of anti-NET antibodies, and impaired NET clearance all associate with disease activity and organ damage in lupus (26). Our group has found something similar in individuals with primary APS (22). Here, we found that high levels of anti-NET IgG and IgM are present in patients hospitalized with COVID-19. Those anti-NET antibodies not only impaired the intrinsic ability of serum DNases to clear NETs, but also associated with impaired respiratory status and overall disease severity. It is possible that these anti-NET antibodies are important orchestrators of an imbalance between NET formation and clearance that perpetuates COVID-19 thromboinflammation.

While the ongoing vaccination campaign is working towards reducing COVID-19 incidence and mortality, millions of survivors of COVID-19 infection continue to suffer from longer-term symptoms of the disease. Certainly, diverse and functional autoantibodies produced during COVID-19 infection are a plausible contributor to the post-COVID syndrome. Intriguingly, one recent study observed that among nine COVID-19 survivors, five developed chronic “long-haul” symptoms and all five had potentially pathological autoantibodies (27). We have previously observed durable anti-NET IgG for up to four years among some APS patients (22). The data presented here suggest the presence of another functional autoantibody in COVID-19, and the persistence and potential long-term consequences of these antibodies warrant further investigation.

## MATERIALS AND METHODS

### Human samples

Serum samples from 328 hospitalized COVID patients were used in this study (**Supplementary Table 1**). Blood was collected into serum separator tubes containing clot activator and serum separator gel by a trained hospital phlebotomist. After completion of biochemical testing ordered by the clinician, the remaining serum was released to the research laboratory. Serum samples were immediately divided into small aliquots and stored at -80°C until the time of testing. All 328 patients had a confirmed COVID-19 diagnosis based on FDA-approved RNA testing. This study complied with all relevant ethical regulations and was approved by the University of Michigan IRB (HUM00179409). Healthy volunteers were recruited through a posted flyer; exclusion criteria for controls included history of a systemic autoimmune disease, active infection, and pregnancy. The 48 controls included 40 females and 8 males, with a mean age and standard deviation of 38 ± 10.

### Human neutrophil purification

Human neutrophils were isolated as we have done and described previously (22).

### Generation of NETs

NETs were generated with PMA stimulation as described previously (28).

### Partial digestion of NETs and quantification of protein

PMA-induced NETs were partially digested with 10 U/ml of Micrococcal nuclease (MNase, NEB) in the presence of Micrococcal nuclease reaction buffer (NEB) for 20 minutes at 37°C. The reaction was stopped by adding Ethylenediaminetetraacetic acid (EDTA, Sigma) to a final concentration of 15 mM. NET protein concentration was determined using Bicinchoninic Acid Kit (Pierce) according to the manufacturer’s instructions.

### Anti-NET IgG and IgM enzyme-linked immunosorbent assays (ELISAs)

A high-binding 96-well EIA/RIA plate (Greiner) was coated overnight at 4°C with MNase-digested NETs diluted to a concentration of 5 µg/ml in 0.05 M bicarbonate buffer (coating buffer). The plate was then washed once with 0.05% Tween 20 (Sigma) in PBS (wash buffer), and blocked with 4% BSA (Sigma) in PBS (blocking buffer) for 2 hours at 37°C. Serum samples were diluted to 1% in blocking buffer, added to the plate, and incubated for 90 minutes at 37°C. The plate was washed 5 times with wash buffer and then incubated for 90 minutes at 37°C with either anti-human IgG-HRP or anti-human IgM-HRP (Jackson Immuno) diluted 1:10000 in blocking buffer. The plate was washed 5 more times with wash buffer and was developed with 3,3′,5,5′-Tetramethylbenzidine (TMB) substrate (Invitrogen). The reaction was stopped by 2N sulfuric acid solution and the absorbance was measured at a wavelength of 450 nm using a Cytation 5 Cell Imaging Multi-Mode Reader (BioTek). Each sample was tested against a corresponding control in which no NETs antigen was plated. This created an individual background value for each sample, which was subtracted to obtain the final result. Schematic illustration of anti-NET ELISA in Figure 1 was created with BioRender.com.

### Purification of human IgG fractions

IgG was purified from COVID-19 or control sera as we have done previously (15).

### Immunofluorescence microscopy

1×10^5^ healthy control neutrophils were seeded onto 0.001% poly-L-lysine coated coverslips as described previously(22). To induce NET formation, neutrophils were incubated in serum-free RPMI media supplemented with L-glutamine and stimulated with 40 nM PMA for 2 hours at 37°C and 5% CO_2_. Following stimulation, cell remnants and NETs were fixed with 4% paraformaldehyde for 10 minutes at room temperature, followed by overnight blocking in 10% fetal bovine serum (FBS) in PBS (blocking buffer). Fixed cells were then incubated with either control or COVID sera (diluted to 10%) for 1 hour at 4°C. NETs were detected by a polyclonal antibody against neutrophil elastase (Abcam Ab21595). IgG was detected by fluorochrome-conjugated DyLight 594 anti-human IgG (Thermo Fisher). Nuclear DNA was detected with Hoechst 33342. Coverslips were mounted with Prolong Gold Antifade (Thermo Fisher) and images were collected with a Cytation 5 Cell Imaging Multi-Mode Reader (BioTek).

### Quantification of S100A8/A9 (calprotectin)

Calprotectin was measured with the Human S100A8/S100A9 Heterodimer DuoSet ELISA (DY8226-05, R&D Systems) according to the manufacturer’s instructions (13).

### Quantification of myeloperoxidase-DNA complexes

Myeloperoxidase-DNA complexes were measured as has been previously described (6, 29).

### NET degradation assay

PMA-stimulated NETs were degraded as previously described with minor modifications (30). Briefly, purified healthy control neutrophils were resuspended in RPMI media (Gibco) supplemented with L-glutamine. Neutrophils (1×10^5^ per well) were then seeded on a 0.001% poly-L-lysine (Sigma)-coated 96-well plate (Costar 3631). To induce NET formation, cells were incubated with 20 nM PMA (Sigma) for 4 hours at 37°C and 5% CO_2_. Following incubation, the culture media was removed and the plate washed gently with PBS. NETs were then stained by incubating the cells with 1 mM SYTOX Green (Thermo Fisher) for 30 minutes at 37°C and 5% CO_2_. The SYTOX solution was gently removed and replaced with PBS, and fluorescence was quantified at excitation and emission wavelengths of 504 nm and 523 nm using a Cytation 5 Cell Imaging Multi-Mode Reader (BioTek). To assess NET degradation, the PBS was gently removed and NETs were incubated for 90 minutes (at 37°C and 5% CO_2_) with COVID serum samples diluted to 5% in nuclease buffer (10 mM Tris-HCl pH 7.5, 10 mM MgCl_2_, 2 mM CaCl_2_, and 50 mM NaCl). In some experiments, NETs were incubated with healthy control serum diluted to 5% in nuclease buffer and supplemented with COVID or control IgG at a final concentration of 500 µg/ml. Each sample was tested in triplicate. MNase (10 U/ml)-treated wells served as the positive control. Following the 90-minute incubation, the serum-containing supernatants were discarded and PBS was added to each well. To quantify residual NETs, SYTOX fluorescence was again measured at excitation and emission wavelengths of 504 nm and 523 nm using a Cytation 5 Cell Imaging Multi-Mode Reader (BioTek). Schematic illustration of NET degradation assay in Figure 4 was created with BioRender.com.

### Statistical analysis

Normally-distributed data were analyzed by 2-sided t test and skewed data were analyzed by Mann-Whitney test. Comparisons of more than two groups were analyzed by one-way ANOVA with correction for multiple comparisons by Dunnett’s method. Correlations were tested by Spearman’s correlation coefficient. Data analysis was with GraphPad Prism software version 8. Statistical significance was defined as p<0.05 unless stated otherwise.

## Data Availability

All data are available in the manuscript and supplement.

## ACKNOWLEDGEMENTS

YZ was supported by career development grants from the Rheumatology Research Foundation, Arthritis National Research Foundation, and APS ACTION. JAM was partially supported by the VA Healthcare System. YK was supported by the Intramural Research Program of the NIH and NHLBI, the Lasker Foundation, the Falk Medical Research Trust Catalyst Award, and the JOBST-American Venous Forum Award. JSK was supported by grants from the NIH (R01HL115138), Lupus Research Alliance, Rheumatology Research Foundation, and Burroughs Wellcome Fund. YK and JSK were supported by the University of Michigan Frankel Cardiovascular Center Ignitor Award and the A. Alfred Taubman Medical Research Institute.

## AUTHORSHIP

YZ, SY, SN, CH, AH, KG, MZ, JAM, and HS conducted experiments and analyzed data. YZ, YK, and JSK conceived the study. All authors participated in writing the manuscript and gave approval before submission.

## Supplementary Materials

**Supplementary Table 1:**
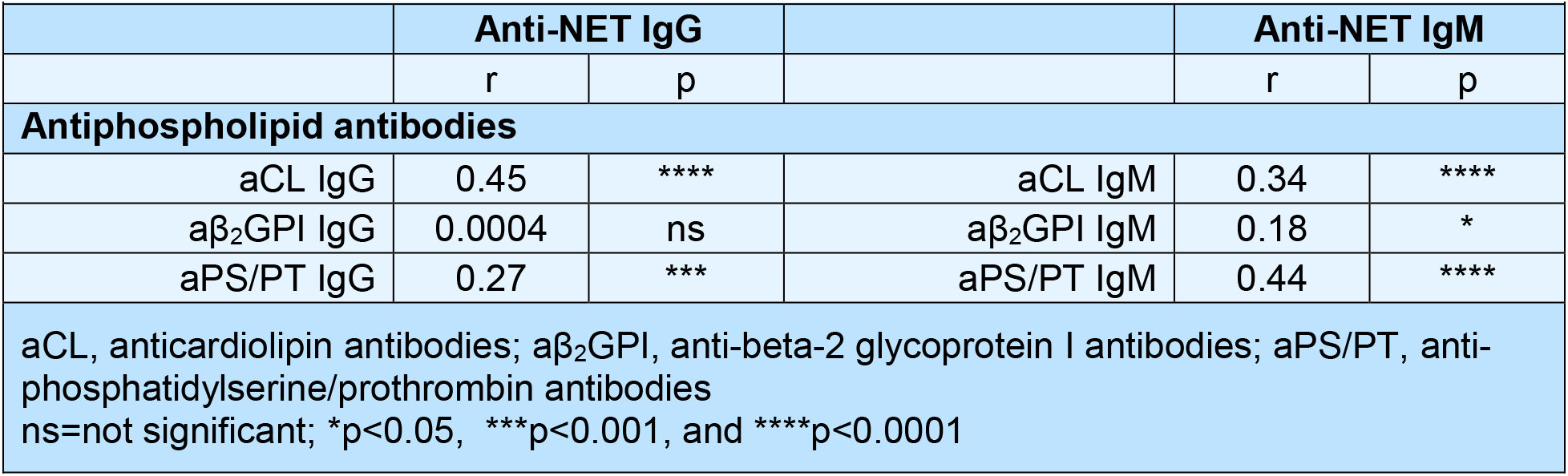
Correlation of anti-NET IgG/IgM with antiphospholipid antibodies in COVID patients

**Supplementary Figure 1:**
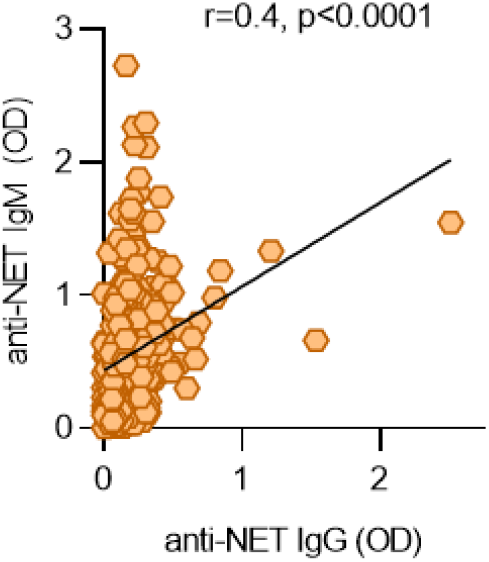
Association of anti-NET IgG with anti-NET IgM. Anti-NET IgG and anti-NET IgM were compared for 328 COVID patients. Spearman’s correlation coefficient was calculated and is shown in the panel.

**Supplementary Figure 2:**
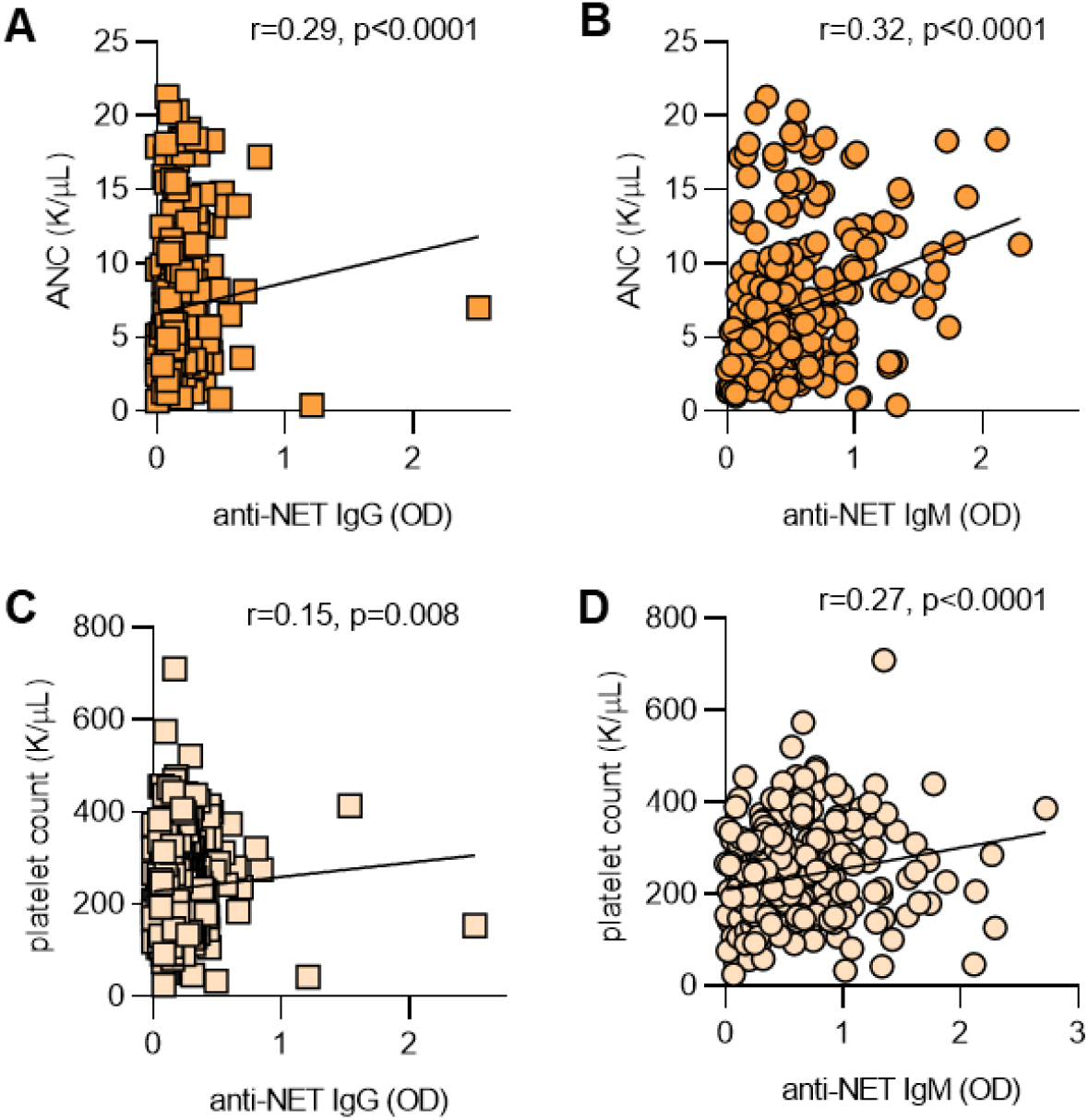
Association of anti-NET IgG/IgM with absolute neutrophil count (ANC) and platelet count. **A-B**, Anti-NET IgG and IgM levels were compared to ANC on the same day as the research sample (n=266 COVID-19 patients). **C-D**, Anti-NET IgG and IgM levels were compared to platelet count on the same day as the research sample (n=315 COVID-19 patients). Spearman’s correlation coefficients were calculated and are shown in the panels. ANC=absolute neutrophil counts.

**Supplementary Figure 3:**
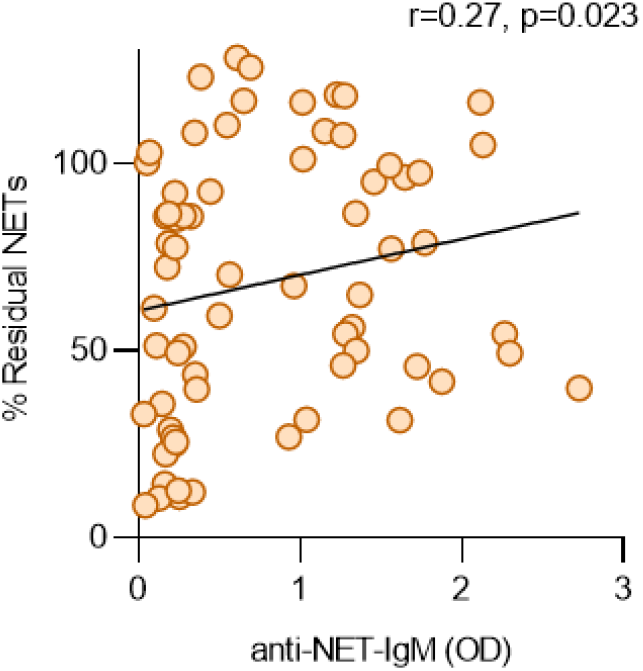
Anti-NET IgM correlates with impaired NET degradation. Freshly-induced NETs were incubated with COVID sera, and percent residual NETs was determined for each sample after 90 minutes. Percent residual NETs was normalized against the mean of untreated NETs. Correlation with anti-NET IgM was determined by Spearman’s method.

